# Evaluation of rapamycin as a neuroprotective treatment in Alzheimer’s disease: a six-month single-arm open-label clinical pilot trial

**DOI:** 10.64898/2025.12.08.25340853

**Authors:** Jonas E. Svensson, Ruben P. Dörfel, Martin Schain, Martin Bolin, Simona Sacuiu, Göran Hagman, Anton Forsberg Morén, Miia Kivipelto, Pontus Plavén-Sigray

## Abstract

**Background:** Rapamycin has demonstrated neuroprotective effects in preclinical Alzheimer’s disease (AD) models, yet clinical data remain limited. Here, we conducted a pilot trial to assess the feasibility and safety of rapamycin treatment in early-stage AD using multimodal neuroimaging and cerebrospinal fluid (CSF) biomarkers.

**Methods:** This single-arm, open-label pilot study enrolled 14 participants with early-stage AD who received oral rapamycin 7 mg weekly for 26 weeks. Thirteen participants completed treatment. The primary outcome was change in cerebral glucose metabolism measured by [^18^F]FDG PET (*K*_*i*_ and SUVR) in pre-specified regions commonly affected in AD: posterior cingulate cortex, precuneus, and temporoparietal cortex. Secondary outcomes included cerebral blood flow (CBF) via ASL MRI, CSF biomarkers, volumetric MRI, and cognition.

**Results:** Rapamycin was well tolerated, with no serious adverse events. No significant metabolic or perfusion changes were observed in primary regions. Exploratory analyses showed increases in [^18^F]FDG SUVR in the putamen, insula, and anterior cingulate cortex, and thalamic CBF. Higher rapamycin blood concentrations correlated with increased [^18^F]FDG SUVR in several regions, including the temporoparietal cortex. CSF analysis demonstrated significant increases in total tau, neurofilament light, A*β*40, and a numerical, non-significant increase in A*β*42 of similar effect size, while p-tau remained largely unchanged, resulting in a significantly decreased p-tau/total tau ratio.

**Conclusions:** This pilot study demonstrates feasibility of rapamycin trials in AD. The primary [^18^F]FDG outcomes showed no decline, and even increases in exploratory regions, in contrast to what would be expected from natural disease progression. The observed elevations in CSF biomarkers warrant further investigation.

## 1 Introduction

Alzheimer’s disease (AD) remains one of the most challenging neurodegenerative disorders, with no current effective disease-modifying treatment in widespread clinical use. Despite decades of research focusing primarily on the amyloid hypothesis, clinical trials targeting amyloid plaques have shown limited success, with persisting concerns regarding their efficacy, safety profiles, and eligibility constraints [1, 2]. This has underscored the need to explore compounds targeting other pathways to achieve disease-modifying effects [3].

Rapamycin (sirolimus) represents a promising candidate in this context [4]. The drug inhibits the mechanistic target of rapamycin (mTOR) protein kinase, which regulates critical cellular processes including cell growth, protein synthesis, and autophagy [5]. Rapamycin, originally developed as an immunomodulator to prevent organ rejection following transplantation, has demonstrated neuroprotective properties in preclinical experiments [3, 4]. Experiments in mouse models of AD have shown that the drug and its analogues can prevent and reverse cognitive deficits [6, 7], reduce amyloid oligomers and tau pathology [8, 9], normalize synaptic plasticity [10], increase cerebral glucose uptake [7], and enhance neurovascular function [11]. It has been proposed that these beneficial effects are mediated by several mechanisms relevant to both normal aging and neurodegeneration, including enhanced cellular autophagy [12, 13, 14], improved cerebral blood flow [15], and bloodbrain barrier protection [15]. Nevertheless, except for an 8-week pilot trial in ten participants [16], no clinical trials have tested the potential therapeutic effect of rapamycin in AD patients.

Here, we aimed to assess the feasibility of running a longer clinical trial of rapamycin treatment in early-stage AD using a broad set of established imaging and CSF biomarkers [17]. We conducted a six-month, single-arm pilot trial in which fourteen participants with early-stage AD received a weekly (intermittent) oral dose of 7 mg rapamycin. The primary outcome was change in cerebral glucose uptake, measured using [^18^F]FDG positron emission tomography (PET). Secondary outcomes included safety (adverse events), change in cerebral blood flow (CBF) using arterial spin labeling (ASL) magnetic resonance imaging (MRI), CSF A*β* and tau markers, and cognitive function.

## 2 Methods

### 2.1 Study design and procedures

This was a single-center, longitudinal, single-arm pilot study assessing rapamycin treatment in patients with early-stage AD. The study was conducted at the Memory Clinic at Karolinska University Hospital Solna in Stockholm, Sweden between September 2023 and January 2025. The study was approved by the Swedish Medical Products Agency (5.1-2023-8283) and the Swedish Ethical Review Authority (2023-03075-02 and 2023-00611-01). Written informed consent was obtained from all participants and their designated study partners before any study procedures were initiated. The trial was registered at clinicaltrials.gov (NCT06022068) and EudraCT (2023-000127-36). The clinical study protocol has been published previously [17].

### 2.2 Recruitment, inclusion and exclusion criteria

Participants were recruited from the Memory Clinic patient population through two pathways: i) patients who had previously consented to be registered in the Geriatric Clinic Database (GEDOC) for clinical research on neurodegenerative diseases, and had recently received an AD diagnosis, and ii) patients who expressed interest after being informed about the trial by their treating physician. Potential participants were pre-screened via phone, and suitable candidates were invited to a screening visit accompanied by a study partner, typically next of kin.

For a full report on inclusion/exclusion criteria, see the published clinical study protocol [17]. In brief, participants were eligible for inclusion if they were between 50 and 80 years of age and were amyloid-positive as assessed by a CSF sample, with a clinical diagnosis of mild cognitive impairment (MCI) or mild dementia of Alzheimer’s type according to the National Institute of Aging-Alzheimer’s Association (NIA-AA) 2018 criteria. For participants with dementia, the disease had to be in an early stage, defined as having a Montreal Cognitive Assessment (MoCA) score ≥ 18. Exclusion criteria included history of major diseases that might have interfered with safe engagement in the intervention (e.g., severe liver or kidney disease, or uncontrolled diabetes), history of major neurological disorders, evidence of clinically relevant psychiatric disorders, contraindications for the use of rapamycin (including medications with known serious interaction risks), significant obesity, untreated hyperlipidemia, and recent use of immunosuppressive medications or experimental medications for AD.

### 2.3 Study visits

Participants were assessed for eligibility during a screening visit, including vital signs, medical history, concomitant medication review, and a physical and neurological examination. Before starting treatment, participants underwent [^18^F]FDG PET imaging to assess brain glucose uptake, and brain MRI to assess cerebral blood flow and grey matter volumetry. The cognitive test scores and CSF data from the diagnostic work-up were used as baseline values for the study. If more than nine months had passed from the last MoCA assessment to the screening, a new MoCA was performed by a neuropsychologist as part of the screening process. For participants where the diagnostic lumbar puncture (LP) was performed more than nine months prior to the screening visit, the protocol allowed for a new LP. However, if the clinical status was judged as stable, older measurements were deemed accepted, since prior research has shown that CSF concentrations of amyloid and tau are relatively stable following diagnosis [18, 19]. Full duration between measurements and start/end of treatment are reported in Supplementary Information 1.

Participants were prescribed oral rapamycin (Rapamune^®^, tablet form) which they collected at the Karolinska University Hospital pharmacy. The initial dose was 3 mg/week, which was increased to the target dose of 7 mg/week from the second week if well tolerated. Throughout the treatment period of 26 weeks, participants were monitored through regular follow-up visits and phone calls: a phone follow-up 1–3 days after the first dose, a clinical visit 5–14 days after the first dose, a second phone follow-up at approximately week 6, and a clinical visit at week 13 that included the collection of blood samples for pharmacokinetic analysis (PK) of the study drug concentration. The results from the collected PK data (whole-blood concentration measured at *C*_min_ and 1 h, 3 h, 48 h post-dose) have been analyzed and reported in a separate publication [20]. In brief, we observed the lowest coefficient of variation in the 48 h sample, and this was thus selected as the primary concentration measure for ensuing statistical analyses described below. At all study visits, adverse events were recorded, and a standard panel of safety blood tests were collected and analyzed (see clinical study protocol for more information [17]).

At the end of the 26-week treatment period, participants underwent follow-up assessments. Follow-up PET examinations were scheduled approximately 14 days after the final rapamycin dose (corresponding to approximately 5 half-lives [20]) to balance potential acute pharmacological effects on CNS glucose metabolism and potential reversion to pre-treatment states. Additionally, cognitive testing was performed and participants were offered a follow-up LP procedure.

### 2.4 Imaging procedures

All brain imaging procedures took place at the Brain Molecular Imaging Center and Magnetic Resonance Center at Karolinska Institutet, Stockholm, Sweden, where participants underwent [^18^F]FDG PET to measure cerebral glucose metabolism, T1-weighted MRI to assess volumetry, and pseudocontinuous ASL MRI to measure cerebral blood flow.

#### 2.4.1 PET

[^18^F]FDG was produced by Karolinska University Hospital according to standard clinical procedures. PET acquisitions were performed on a GE Discovery MI 5 PET/CT system. Each acquisition was preceded by a low dose computed tomography (CT) scan for attenuation correction. At the start of the PET acquisition, participants received an intravenous bolus injectionof [^18^F]FDG in the antecubital vein (mean 156 ± 33 SD MBq for the baseline scan, mean 157 ± 35 SD MBq for the follow-up), followed by an injection of approximately 10 mL of saline.

During the first 10 minutes post injection, emission data was acquired over the participant’s thorax to estimate an image-derived input function. This data was reconstructed into 27 timeframes (2 mm × 2 mm × 2.8 mm voxels) with the following frame definition: 10 × 5 s, 4 × 10 s, 2 × 15 s, 3 × 20 s, 2 × 30 s, 6 × 60 s, without motion correction, using TOF-OSEM with 68 updates (two iterations, 34 subsets) and all conventional PET corrections applied.

Emission data for the brain was subsequently acquired between 30- and 60-minutes post injection, and reconstructed into 10 time-frames (1 mm × 1 mm × 2.8 mm voxels), with the following frame definition: 5 × 120 s, 5 × 240 s. To correct for motion, a data-driven procedure was used [21]. In brief, list-mode data were binned with fixed frame counts followed by reconstruction into short frames using four iterations of time-of-flight maximum-likelihood expectation maximization (TOF-MLEM) into 4 mm × 4 mm × 2.8 mm voxels without point spread function, attenuation, or scatter corrections. Motion was estimated by rigidly registering the short frames to a common reference frame, which in turn was co-registered to the attenuation correction CT. Estimated motion parameters were incorporated as event-level corrections during the main list-mode reconstruction in addition to all other PET corrections. The reconstruction algorithm used time-of-flight ordered-subsets expectation maximization (TOF-OSEM) with 224 updates (eight iterations, 16–112 subsets).

For each examination, a total of 5 venous blood samples were collected to measure whole-blood and plasma concentrations of [^18^F]FDG radioactivity, at 20, 35, 45, 60, and 90 minutes post radioligand injection.

#### 2.4.2 MRI

All MR acquisitions were performed on a 3 T Signa Premier MRI scanner using a 48 channel head coil (GE Medical Systems, Chicago, Illinois, USA). ASL-MR images were acquired using the GE product 3D pseudo-continuous sequence with post-labeling delays (PLD) of 1.525 and 2.525 s. Quantitative images of cerebral blood flow (CBF) were calculated automatically on the scanner by vendor-provided software. Finally, a high-resolution three-dimensional T1-weighted structural image was acquired using a sagittal, magnetization-prepared rapid gradient echo sequence.

### 2.5 Other assessments

All study participants underwent an LP as part of the routine clinical work-up prior to the diagnosis of AD (i.e., before the inclusion date) and were asked to perform a second LP within 28 days after the last dose of the study drug. CSF was collected in sterile polypropylene tubes and analyzed at Karolinska University Hospital Laboratory where amyloid-*β* (A*β*) 40, A*β*42, phosphorylated tau 181 (p-tau), and total tau were measured with the Lumipulse G-series (Fujirebio Europe N.V., Gent, Belgium) fully automated chemiluminescent enzyme immunoassay [22]. Neurofilament light chain (NfL) and albumin concentrations were also part of the LP-panel. In addition, we report the ratio between A*β*42/40 and CSF-albumin/plasma-albumin. As a post-hoc analysis, we assessed the change in p-tau/t-tau which is not part of routine diagnostic reporting but has been proposed as a complementary marker reflecting the relative burden of phosphorylated tau species, and considered more closely linked to neurotoxicity relative to total tau pathology [23, 24].

Each participant underwent cognitive testing, performed by a trained neuropsychologist, including MoCA, the Rey Auditory Verbal Learning Test, the Rey Complex Figure and Hagman Tests A and B.

### 2.6 Data analysis

In addition to the study protocol pre-registered at clinicaltrials.gov (NCT06022068), the image analysis procedure and statistical analysis plan (SAP) were pre-registered at aspredicted.org (https://aspredicted.org/vz84-t4b6.pdf).

#### 2.6.1 MRI data

T1-weighted MRI images were processed using FastSurfer [25] to segment cortical and subcortical brain regions. Subcortical structures were defined using the Automated Segmentation atlas [26], and cortical parcels were delineated using the Desikan-Killiany atlas [27]. Pre-registered primary regions of interest (ROIs) for [^18^F]FDG PET analysis were the posterior cingulate cortex, precuneus, and temporoparietal lobe. We restricted the primary analysis to these three regions because hypometabolism in these areas is a well-established early feature of AD [28, 29], and limiting the number of statistical tests minimizes the risk of type I errors. Exploratory analyses included the putamen, caudate, anterior cingulate cortex (ACC), insula, hippocampus, amygdala, frontal cortex, occipital cortex and total cerebral grey matter.

For the ASL measurements, the T1-weighted images were co-registered to the M0 scan using boundary-based registration [30]. The FastSurfer segmentation masks were applied to the ASL data to extract median regional cerebral blood flow values from the PLD 1.525 s and 2.525 s images, respectively. For visualization purposes, all individual images were also registered to a standard stereotactic space (the MNI152NLin2009cAsym template) using a non-linear transformation [31].

#### 2.6.2 PET data

The input functions for the quantification of brain PET data were derived following a procedure described in detail elsewhere [32]. In brief, aortic masks were segmented on thorax CT attenuation maps using TotalSegmentor [33], followed by morphological erosion (eight voxels). The masks were then applied to the thorax PET scans to extract image-derived input functions reflecting [^18^F]FDG concentrations in arterial blood during the first 10 minutes post-injection. The image-derived input functions were then concatenated with manual venous plasma samples to derive complete input functions from time of injection to the end of the scan session.

Segmentations obtained from FastSurfer were registered and projected to the brain PET images to derive regional time-activity curves (TACs). Patlak Graphical Analysis was thereafter used to estimate the net influx rates of [^18^F]FDG (*K*_*i*_) for each brain region [34].

Additionally, regional [^18^F]FDG standardized uptake value ratios (SUVR) were calculated by dividing the area under each regional TAC by that of the cerebellum, as this region traditionally has been considered a “pathologically preserved” brain region, and is commonly used as a reference region for PET studies in AD (see Supplementary Information 2 for construction of reference region masks). The ensuing cerebellar reference region showed negligible difference between follow-up and baseline in *K*_*i*_ (2.4 ± 12.0%, p = 0.75) or SUV area under the curve (-1.0 ± 10.2%, p = 0.70) estimates, suggesting the [^18^F]FDG uptake is stable in this region before and after treatment.

### 2.7 Statistical analysis

Change between baseline and follow-up for regional [^18^F]FDG *K*_*i*_ and SUVR values were assessed using a linear mixed effect model with timepoint as independent variable, ROI×timepoint as interaction effect, and subject modelled as varying intercepts, supplemented by post-hoc paired t-tests for each ROI. Regional CBF differences between baseline and follow-up were assessed using a linear mixed effect model, with CBF estimates at PLD 1.525 s and 2.525 s as dependent variables, timepoint and a binary PLD-indicator as fixed effects, and subject-ID as random effect. Changes in cognitive test scores and CSF markers before and after treatment were analyzed using paired t-tests. The significance level for all tests was set at 0.05.

In addition, we performed a set of exploratory analyses to further investigate potential treatment-related effects. We examined whether rapamycin exposure was related to study outcomes by correlating whole-blood rapamycin concentration at 48 hours post-dosing with changes in regional [^18^F]FDG PET, cerebral blood flow, CSF markers and MoCA using linear regression. As an additional post-hoc analysis, we explored associations between changes in brain volumetric measures, [^18^F]FDG uptake, cerebral blood flow and MoCA and changes in CSF markers using linear regression.

### 2.8 Deviation from pre-registered data analysis plan

The primary endpoint defined in the preregistered data analysis plan of this study was the within subject change in metabolic rate of glucose (MRGlu). MRGlu is derived by multiplying the [^18^F]FDG *K*_*i*_ values with plasma glucose concentration and dividing by a lumped constant. During the course of this study, the equipment used for glucose measurements at the PET examinations malfunctioned, resulting in unreliable measurements. We instead report the [^18^F]FDG *K*_*i*_ values, rather than MRGlu, alongside SUVR estimates, as our primary endpoint. To assess the longitudinal stability of plasma glucose within participants, fasting glucose levels obtained as part of the safety assessments and measured at Karolinska University Hospital central laboratory were analyzed. From these measurements, we found that the mean absolute percentage variability of plasma glucose over the treatment period was less than 4% (3.82% ± 2.2% SD). No significant differences were observed between plasma glucose concentrations measured at baseline and those values measured closest to follow-up PET examinations (p = 0.41), indicating that glucose values were stable from baseline to follow-up assessments. In addition, [^18^F]FDG SUVR values, which largely mitigate the influence of plasma glucose by normalizing uptake in target regions to the uptake in cerebellum, showed a significant positive correlation to *K*_*i*_ estimates (see Supplementary Information sFigure 1). These findings support the use of change in [^18^F]FDG *K*_*i*_ and SUVR values as a substitute for change in MRGlu in this longitudinal study.

**Figure 1.**
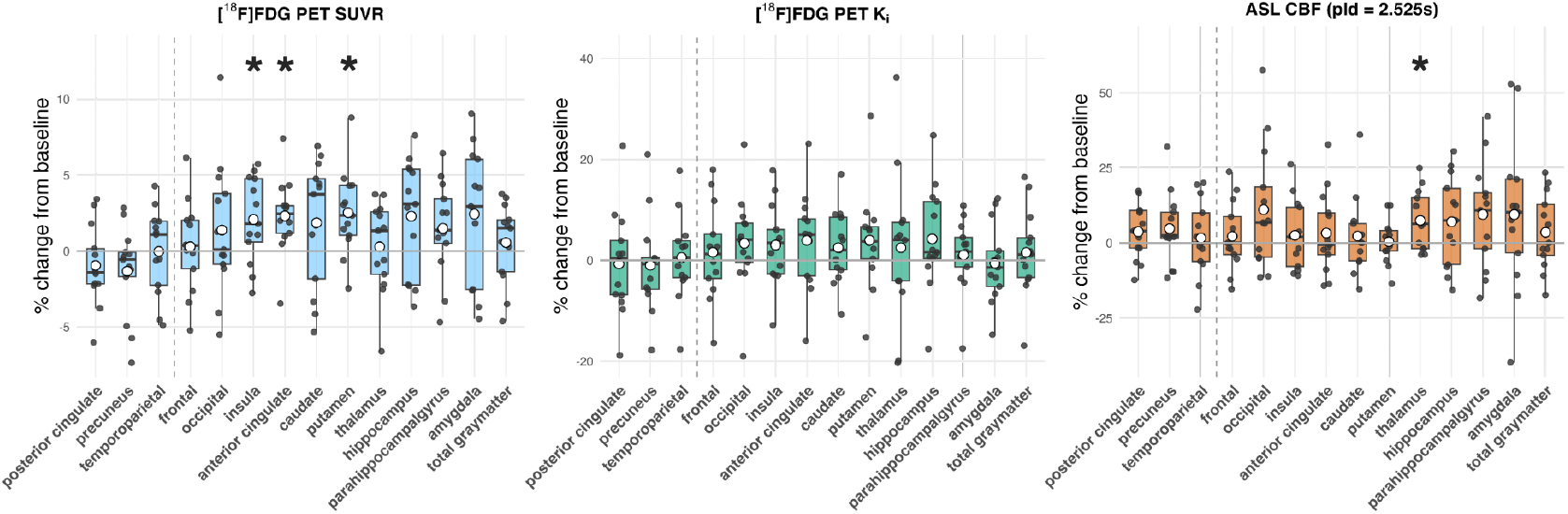
Regional changes from pre-treatment to post-treatment in [^18^F]FDG PET SUVR, [^18^F]FDG PET *K*_*i*_, and MRI ASL CBF (PLD = 2.525 s). No significant changes were observed in the pre-registered primary ROIs (left of vertical dashed lines) following rapamycin treatment. The exploratory analyses showed significant increases in SUVR in the putamen, insula, and anterior cingulate cortex, as well as increased CBF in the thalamus following treatment.

## 3 Results

### 3.1 Participant characteristics

Twenty-one patients were contacted through a pre-screening telephone call. Of these, 14 patients attended an in-person screening visit, and all were enrolled in the study. One male participant discontinued the study within two weeks of initiating treatment due to intolerance of the study drug (nausea), resulting in 13 participants completing the 26-week treatment period. All participants were on stable doses of cholinesterase inhibitors for at least 4 weeks prior to initiation of the study drug. Baseline characteristics for all enrolled participants are presented in Table 1.

**Table 1:**
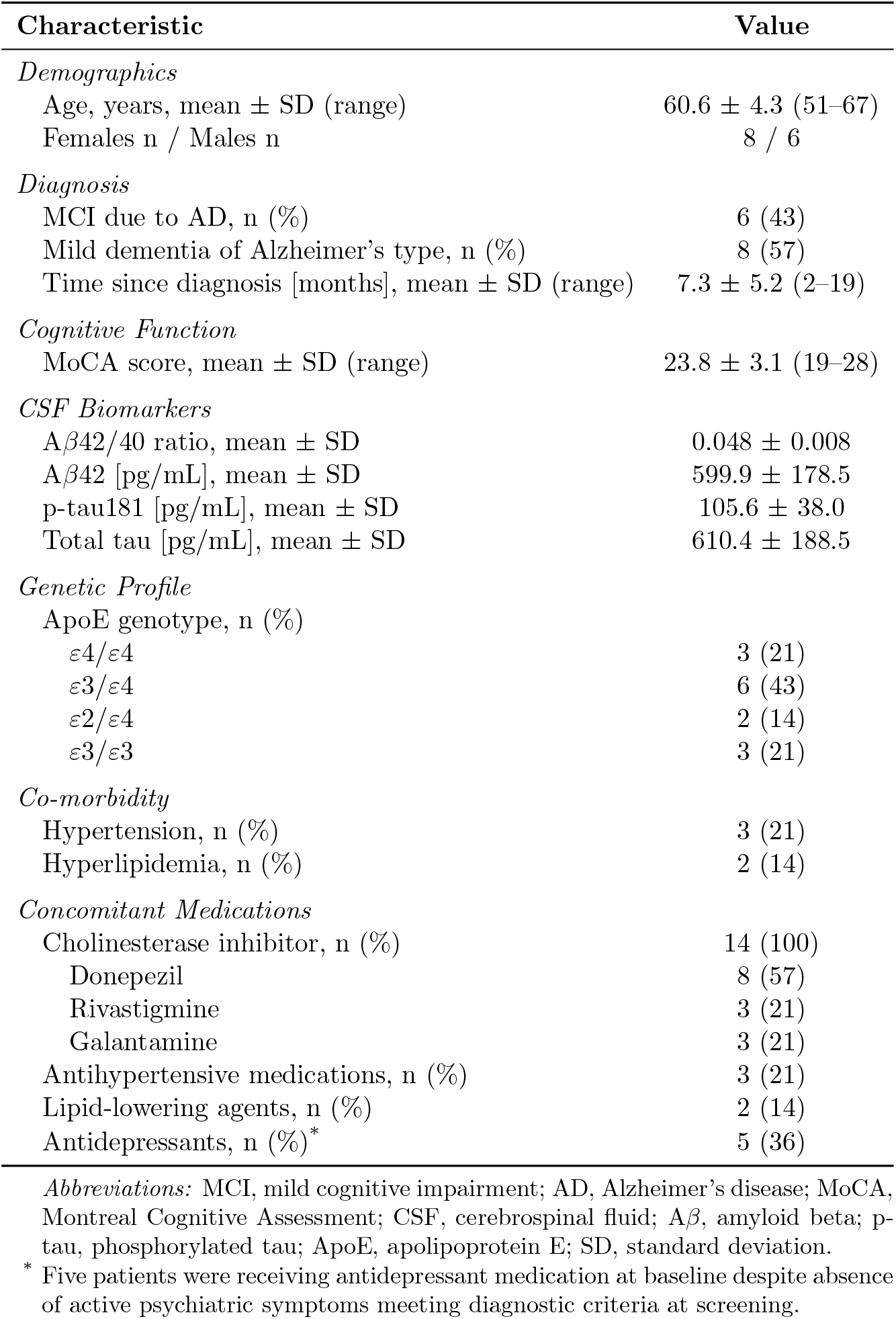
Baseline characteristics of all enrolled study participants (N = 14)

| Characteristic | Value |
| --- | --- |
| *Demographics* |  |
| Age, years, mean ± SD (range) | 60.6 ± 4.3 (51–67) |
| Females n / Males n | 8 / 6 |
| *Diagnosis* |  |
| MCI due to AD, n (%) | 6 (43) |
| Mild dementia of Alzheimer’s type, n (%) | 8 (57) |
| Time since diagnosis [months], mean ± SD (range) | 7.3 ± 5.2 (2–19) |
| *Cognitive Function* |  |
| MoCA score, mean ± SD (range) | 23.8 ± 3.1 (19–28) |
| *CSF Biomarkers* |  |
| Aβ42/40 ratio, mean ± SD | 0.048 ± 0.008 |
| Aβ42 [pg/mL], mean ± SD | 599.9 ± 178.5 |
| p-tau181 [pg/mL], mean ± SD | 105.6 ± 38.0 |
| Total tau [pg/mL], mean ± SD | 610.4 ± 188.5 |
| *Genetic Profile* |  |
| ApoE genotype, n (%) |  |
| ε4/ε4 | 3 (21) |
| ε3/ε4 | 6 (43) |
| ε2/ε4 | 2 (14) |
| ε3/ε3 | 3 (21) |
| *Co-morbidity* |  |
| Hypertension, n (%) | 3 (21) |
| Hyperlipidemia, n (%) | 2 (14) |
| *Concomitant Medications* |  |
| Cholinesterase inhibitor, n (%) | 14 (100) |
| Donepezil | 8 (57) |
| Rivastigmine | 3 (21) |
| Galantamine | 3 (21) |
| Antihypertensive medications, n (%) | 3 (21) |
| Lipid-lowering agents, n (%) | 2 (14) |
| Antidepressants, n (%)<sup>*</sup> | 5 (36) |

*Abbreviations:* MCI, mild cognitive impairment; AD, Alzheimer’s disease; MoCA, Montreal Cognitive Assessment; CSF, cerebrospinal fluid; A $\beta$ , amyloid beta; p-tau, phosphorylated tau; ApoE, apolipoprotein E; SD, standard deviation.
<sup>\*</sup> Five patients were receiving antidepressant medication at baseline despite absence of active psychiatric symptoms meeting diagnostic criteria at screening.

### 3.2 Dosing and treatment adherence

Of the 13 participants who completed the study, 11 (85%) received the target dose of 7 mg weekly rapamycin. Two participants (15%) initially received reduced doses due to safety considerations or side effects: one participant received 2 mg weekly due to vertigo following the initial dose, and another received 4 mg weekly due to idiopathic chronically elevated aspartate transaminase levels.

Both of these participants were able to tolerate dose increases later in the study (but after the collection of PK samples), with an increase of 2 to 4 mg and 4 to 7 mg after the 13-week follow-up, respectively. Treatment adherence was verified during clinical follow-up visits via inspection of dosette boxes. Temporary treatment interruptions (one week) occurred in two participants due to infection; otherwise, all participants adhered to the dosing schedule.

### 3.3 Safety and tolerability

No serious adverse events were reported during the trial. The most commonly reported adverse events were gastrointestinal symptoms, which included episodes of diarrhea (6/14 participants, 43%), and nausea (2/14 participants, 14%). Three participants (21%) experienced mouth sores, with recurring episodes for two participants. Additionally, two participants (14%) experienced bacterial infections (erythema migrans and a tooth infection), requiring treatment with an antibiotic. Two participants reported upper respiratory tract infections. All adverse events were either mild (94%) or moderate (6%) in severity and were resolved spontaneously, following dose reduction, or discontinuation of the drug. Safety laboratory assessment of blood samples showed no clinically significant alterations throughout the study.

### 3.4 Primary outcome – [^18^F]FDG PET

The primary endpoint analysis showed no statistically significant changes in [^18^F]FDG uptake following 26 weeks of rapamycin treatment (Figure 1). For the pre-specified primary regions of interest, *K*_*i*_ values were not significantly different between baseline and follow-up: posterior cingulate cortex (-0.7 ± 10.4%, p = 0.62), precuneus (-1.0 ± 9.7%, p = 0.60), and temporoparietal lobe (+0.6 ± 8.6%, p = 0.97) (Figure 2). Similarly, SUVR measurements using cerebellar cortex as reference showed no significant changes in the primary regions (posterior cingulate: -0.9 ± 2.6%, p = 0.23; precuneus: -1.3 ± 3.0%, p = 0.17; temporoparietal lobe: -0.03 ± 2.9%, p = 0.99) (Figure 2). Average pre-post treatment brain images for [^18^F]FDG PET *K*_*i*_ and SUVR values are shown in Figure 2.

**Figure 2.**
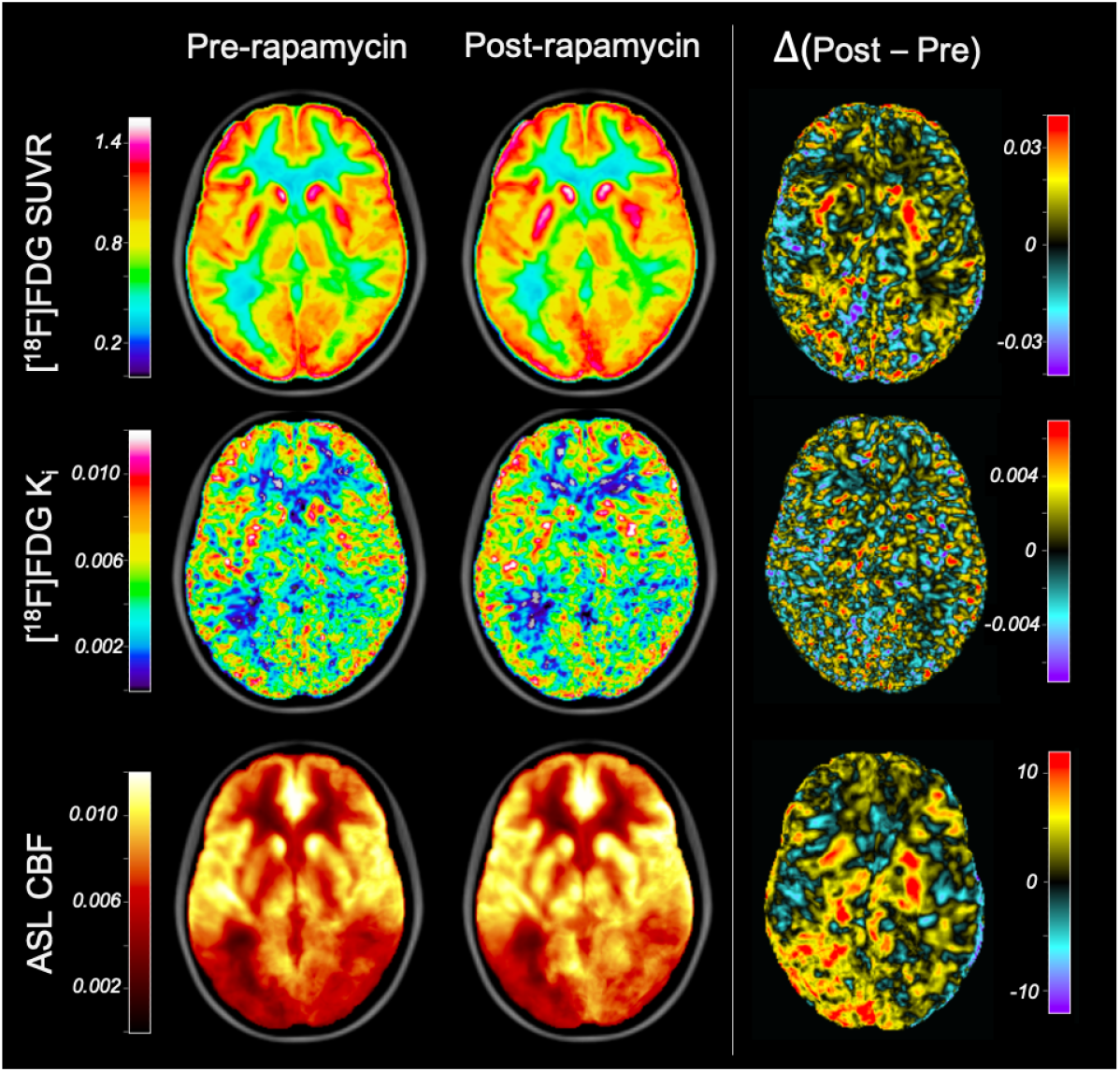
Average [^18^F]FDG PET (SUVR and *K*_*i*_) and MRI ASL (cerebral blood flow) images before and after treatment, along with corresponding average difference maps (post treatment – pre treatment).

In the exploratory analyses of additional brain regions (see Supplementary Information sTable 1 and sTable 2), the largest change in uptake was observed for [^18^F]FDG SUVR in the putamen (+2.5 ± 2.8%, p = 0.006), insula (+2.1 ± 2.8%, p = 0.024), and ACC (+2.3 ± 2.5%, p = 0.008). Following a pre-registered secondary analysis, we observed a significant positive correlation between the whole-blood concentration of rapamycin and the change in [^18^F]FDG SUVR in multiple brain regions (Figure 3): temporoparietal cortex (r = 0.68, p = 0.02), amygdala (r = 0.69, p = 0.02), insula (r = 0.67, p = 0.02), hippocampus (r = 0.75, p = 0.01) and parahippocampal gyrus (r = 0.65, p = 0.03).

**Figure 3.**
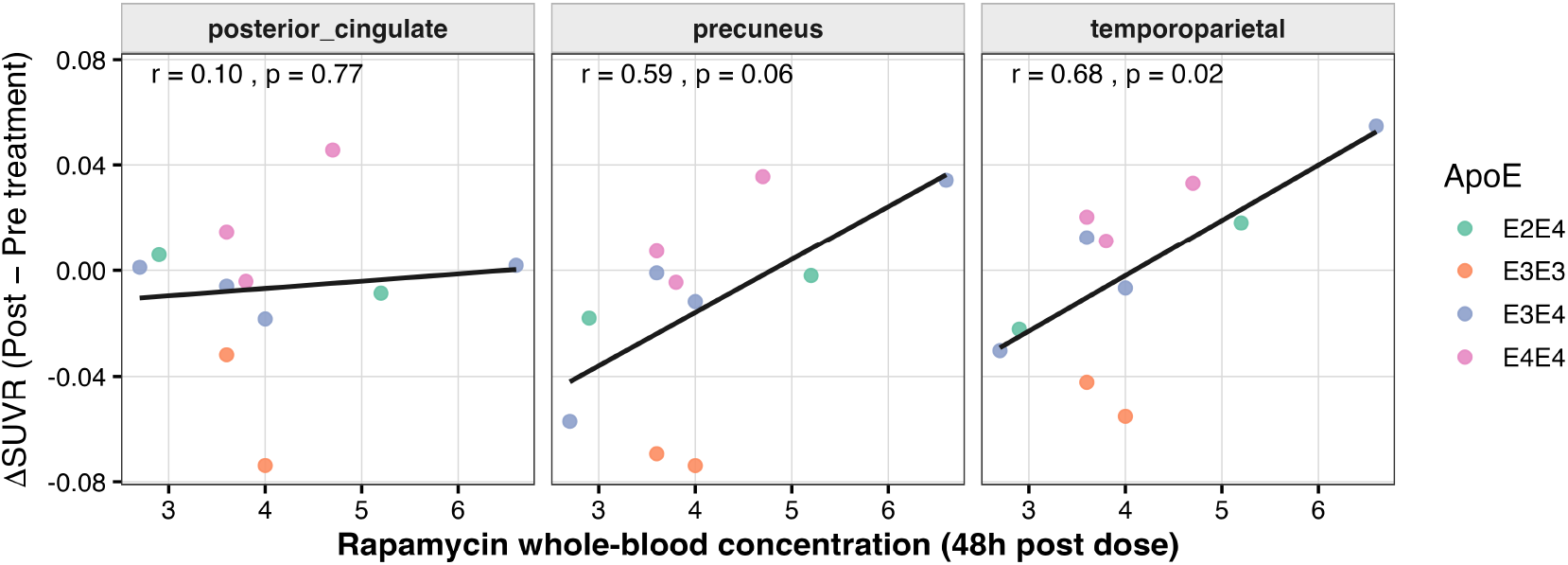
Dose-response association between change in [^18^F]FDG PET SUVR and blood concentration of rapamycin (ng/mL) at the 48 h PK-sample taken approximately at week 13 into the treatment in the primary regions of interest. Two subjects increased their dose of rapamycin (from 2/4 mg to 4/7 mg, respectively) after this timepoint and have therefore been excluded from this analysis.

### 3.5 Secondary and exploratory outcomes

#### 3.5.1 Cerebral Blood Flow

ASL MRI measurements acquired using two post-labeling delays (1.525 s and 2.525 s) showed no significant changes in regional CBF in the primary regions of interest (Figure 1). The linear mixed-effects model showed no significant effect of timepoint (pre-vs. post-treatment) on CBF estimates (all p > 0.18), with mean ± SD changes ranging from -2.0 ± 19.8% to 4.8 ± 12.0% across all three primary ROIs for both PLDs (1.525 s and 2.525 s, respectively). The interaction between timepoint and PLD was not significant (all p > 0.40), indicating consistent results across both PLD protocols. There were no significant CBF changes in any of the exploratory regions for the PLD protocols (Supplementary Information sTable 3 and sTable 4), except for the thalamus (+7.5 ± 9.7%, p = 0.02). There were no significant correlations between rapamycin blood concentration at 48 h post-dosing and change (p > 0.39 for both PLDs).

#### 3.5.2 Volumetric measures

Structural MRI analysis revealed significant gray matter volume reductions consistent with expected disease progression. Total gray matter volume decreased by an average of 1.2 ± 1.5 SD% over the six-month treatment period (p = 0.013). Significant volume reductions were observed in all primary regions: temporoparietal (-1.8 ± 1.9%, p = 0.002), precuneus (-1.7 ± 2.9%, p = 0.043), and posterior cingulate (-1.3 ± 2.4%, p = 0.035).

Secondary regions, such as the hippocampus (-1.2 ± 2.0%, p = 0.044), showed similar patterns of volume reduction, though not all were statistically significant (see Supplementary Information sTable 5). There was no significant correlation between rapamycin blood concentration at 48 h post dose and change in volume for any of the primary regions (all p > 0.16).

#### 3.5.3 CSF markers

Analysis of CSF biomarkers for participants with both baseline and follow up LP (N = 10, see Supplementary Information 1 and sTable 6 and sFigure 2) demonstrated increases in concentration across multiple proteins following rapamycin treatment (Figure 4): total tau increased by an average of 21.8 ± 16.7 SD% (p = 0.003), while NfL showed a 23.5 ± 26.0% increase (p = 0.01). p-tau181 showed no significant change (4.5 ± 16.9%, p = 0.59), while the p-tau181/total tau ratio decreased by -14.1 ± 8.0% (p = 0.001).

**Figure 4.**
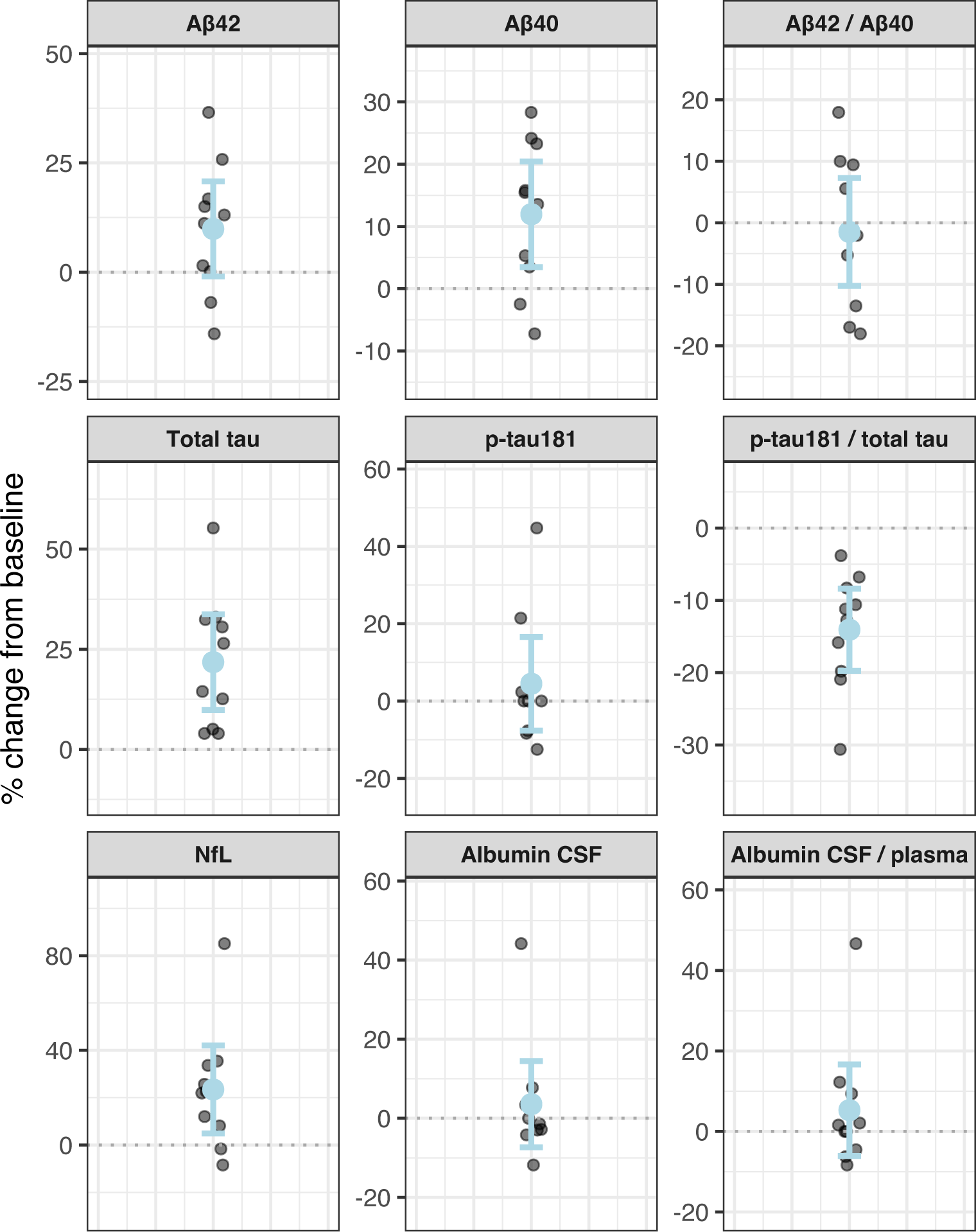
Percentage change from preto post-treatment in cerebrospinal fluid (CSF) biomarkers, shown as individual delta values with group mean and 95% confidence intervals. Markers include A*β*42, A*β*40, A*β*42/40 ratio, total tau, p-tau181, p-tau181/total tau ratio, NfL, CSF albumin, and the CSF/plasma albumin ratio.

A*β* levels also showed elevations, with A*β*40 increasing by 12.0 ± 11.9% (p = 0.006) and A*β*42 showing a 9.9 ± 15.2% numerical but non-significant increase (p = 0.18). The A*β*42/40 ratio remained stable (-1.5 ± 12.3%, p = 0.57). CSF albumin levels and the CSF/plasma albumin ratio showed no significant changes (3.6 ± 15.2%, p = 0.43 and 5.3 ± 15.9%, p = 0.34, respectively).

There was no significant correlation between change in regional volumes (as assessed using a T1-weighted MR protocol) and change in any of the CSF markers following treatment (see Supplementary Information sTable 7). There were no significant correlations between rapamycin blood concentration at 48 h post dose and change in any of the CSF markers (all p > 0.14).

#### 3.5.4 Cognitive outcomes

MoCA scores remained stable from baseline to follow-up, with a mean ± SD % change of +1.1 ± 9.8 (p = 0.54). Individual cognitive domain assessments, including Rey Auditory Verbal Learning Test, Rey Complex Figure Test and Hagman test A and B, showed no significant changes (all p > 0.14) (see Supplementary Information sTable 8). There was no significant association between rapamycin blood concentration at 48 h post-dose and pre-post treatment change in MoCA scores (r = 0.14, p = 0.71).

## 4 Discussion

This single-arm pilot study represents one of the first clinical trials evaluating rapamycin as a potential neuroprotective treatment in patients with early-stage Alzheimer’s disease. The study demonstrates the feasibility—including regulatory approval, patient recruitment, safety, and the collection of neuroimaging and fluid biomarkers—of conducting a larger repurposing trial of rapamycin in AD. The primary outcome measure, cerebral glucose metabolism measured through [^18^F]FDG PET uptake in the posterior cingulate cortex, precuneus, and temporoparietal cortex did not change following treatment. However, exploratory analyses showed regional increases in [^18^F]FDG PET SUVR, most notably in the putamen, insula, and anterior cingulate cortex. Rapamycin blood concentrations at 48 hours post-dosing were positively associated with changes in [^18^F]FDG uptake in several brain regions, suggesting a potential dose–response relationship. Cerebral blood flow measured by ASL MRI remained largely stable, except for an increase in thalamic perfusion. Analysis of CSF biomarkers demonstrated increases in A*β*40, NfL, and total tau, whereas p-tau remained unchanged, leading to a significant decrease in the p-tau/total tau ratio.

### 4.1 Neuroimaging

For the pre-specified primary regions of interest (posterior cingulate cortex, precuneus, and temporoparietal lobe), [^18^F]FDG *K*_*i*_ and SUVR estimates showed no significant changes. However, increased glucose uptake was observed in several exploratory brain regions. No significant decreases were observed in any of the analyzed brain regions.

ASL MRI measurements demonstrated stable cerebral blood flow throughout the treatment period. The linear mixed-effects model showed no significant effect of timepoint on CBF values, with mean regional changes ranging from -2% to 5% in our primary regions. A previous study in healthy APOE4 carriers reported that rapamycin treatment increased CBF by >15% in multiple areas, such as total gray matter and precuneus [35]. When excluding the non-APOE4 carriers and reanalyzing the data from our trial (n = 11), we could not reproduce these effect sizes (data not shown). Analysis of structural MRI showed progressive gray matter atrophy (see Supplementary Information sTable 5), consistent with literature reports of approximately 2% annual gray matter decline in AD [36]. Yet, the observed hippocampal volume loss of 1.1% over six months appears numerically lower than the reported annual decline of 4–5% [37, 38].

Previous literature reports progressive [^18^F]FDG and cerebral blood flow [39] decline in AD patients [40, 41], with even greater decline rates among APOE4 carriers [42, 43, 44]. The absence of significant decreases observed in our primary regions, together with regional increases observed in exploratory analyses, is noteworthy since it does not follow the expected trajectory of natural disease progression and is compatible with a neuroprotective effect. Yet, a control arm and larger sample size would be needed to make strong inference of such effects.

### 4.2 CSF markers

Analysis of CSF biomarkers showed significant increases in NfL, total tau, and A*β*40, and a numerical, but non-significant increase of similar effect size in A*β*42. These findings parallel those reported in the only other published AD-rapamycin trial by Gonzales et al. [16], who demonstrated broad increases in CSF markers (A*β*42, A*β*40, p-tau and NfL) after only 8 weeks of daily rapamycin treatment. The short duration of that trial suggests that the shifts in CSF markers are unlikely to reflect natural disease progression and instead represent a reproducible drug effect. A difference between our study and that of Gonzales et al. is the reduction in the p-tau/total tau ratio observed
here; their exploratory analyses showed increases in phosphorylated tau and in epitope-matched total tau, resulting in no significant change in p-tau/total tau ratios [16]. There are multiple potential explanations for the shifts observed in CSF markers, such as: (i) enhanced autophagy-mediated protein clearance from neurons (i.e., secretory autophagy) [45, 46], (ii) reduction of CSF turn-over [47, 48], and/or (iii) detrimental effects on neural cells [49, 50]. There were no significant correlations between increases in total tau or NfL and loss of gray matter volume (assessed using MRI) in any of the regions included in our study, implying that the observed increases in CSF are not related to acute accelerated atrophy. However, several factors complicate the interpretation of CSF results in our trial: baseline LP was performed during diagnostic clinical workup (median 8 months prior to enrollment, see Supplementary Information 1), and all participants started cholinesterase inhibitors during the same time period. Still, the CSF results reported here and by Gonzales et al. warrant further investigation.

### 4.3 Safety and tolerability

Rapamycin demonstrated an acceptable safety profile in this AD population, with no serious adverse events reported. The most common adverse events were mild gastrointestinal symptoms and mouth sores, consistent with rapamycin’s established side effects. All safety parameters remained within clinically acceptable ranges, including stable HbA1c and systolic blood pressure. This contrasts with data reported by Gonzales et al. [16], who showed small but significant increases in both these parameters. The discrepancy may reflect population differences, dosing regimens (weekly vs. daily), or chance findings in small samples. While the immediate tolerability supports the feasibility of larger rapamycin trials in AD, the observed elevated CSF biomarkers warrant caution and careful monitoring until the mechanism behind the shifts has been better understood.

### 4.4 Limitations

The single-arm design constrains interpretation of observed changes, preventing a clear distinction between treatment effects, natural disease progression, and regression towards the mean. The small sample size (n = 13) also limits the statistical power to detect modest but potentially meaningful changes in the study outcomes. The long interval between baseline assessments (CSF markers and cognition) and treatment initiation complicates interpretation of these data, particularly since all participants initiated treatment with cholinesterase inhibitors during this period. Technical challenges included the collection of reliable plasma glucose measurements, necessitating the use of *K*_*i*_ and SUVR rather than MRGlu as [^18^F]FDG outcome measures. Follow-up PET imaging was scheduled after approximately five rapamycin half-lives to reduce the risk of capturing possible acute pharmacological effects on neuronal glucose uptake rather than persistent treatment-related changes. While this approach was intended to strengthen interpretability, it also reduces the possibility to detect short-lived or transient drug effects. Together, these limitations underscore the pilot nature of this study and emphasize that the presented findings should be considered hypothesis-generating rather than definitive.

## 5 Conclusions

This pilot study demonstrates the feasibility of conducting a clinical trial of rapamycin in early-stage AD. Neither our study nor a previous similar trial showed improvements in primary efficacy measures, though both documented consistent elevations in CSF biomarkers that warrant further investigation. In the pre-registered primary regions of interest, [^18^F]FDG PET and MRI CBF outcomes remained stable over six months. Notably, several exploratory regions showed increases in [^18^F]FDG SUVR, a pattern that could be compatible with a neuroprotective effect of the study drug. The absence of decreases in metabolic and perfusion measures, despite the expected decline in this patient population, may in itself suggest relative preservation of function, though such a conclusion requires confirmation in larger controlled trials.

## Data Availability

All data produced in the present study are available upon reasonable request to the authors

## Acknowledgements

We would like to thank Lars Farde for his valuable feedback on the study design. The study was supported by a Longevity Impetus grant from the Norn Group, Åhlén Stiftelsen, Demensfonden, The Swedish Society of Medicine (SLS), Åke Wibergs Stiftelse (M24-0117), Loo and Hans Osterman Stiftelse, Stiftelsen för Ålderssjukdomar Karolinska Institutet, Stiftelsen för Gamla Tjänarinnor, Tore Nilssons Stiftelse för Medicinsk Forskning, Magnus Bergvalls stiftelse, Karolinska Institutet Research Grants, Stiftelsen Stockholms Sjukhem, Region Stockholm (ALF grant), The Swedish Brain Foundation (PS2021-0012 and PD2024-0444-HK-155), and KI CIMED. None of the funding bodies had any role in the design of the study or in writing the manuscript.

## Authors’ contributions

JES: Conceptualization, Methodology, Software, Formal analysis, Investigation, Resources, Writing – original draft, Supervision, Project administration, and Funding acquisition. RPD: Methodology, Software, Formal analysis, Data curation, Writing – original draft. MS: Methodology, Writing – original draft. MB: Methodology, Formal analysis, Writing – review & editing. SS: Investigation, Writing – review & editing. GH: Investigation, Writing – review & editing. AFM: Resources, Writing – review & editing. MK: Conceptualization, Resources, Supervision, Writing – review & editing. PP-S: Conceptualization, Methodology, Software, Formal analysis, Resources, Data curation, Writing – original draft, Supervision, Project administration and Funding acquisition. All authors have read and approved the final manuscript.

