## Supplemental information for "Evaluation of rapamycin as a neuroprotective treatment in Alzheimer’s disease: a six-month single-arm open-label clinical pilot trial"

### Supplementary information

#### Supplementary information 1

The median duration between the baseline [18F]FDG scan and the first dose of rapamycin was 15 days (sd = 12.7), and the median duration between the last dose and follow-up scan was 14 days (sd = 2.4). The median duration between the first LP and the first dose of rapamycin was 260 days (sd = 281). 10 subjects consented to perform a follow-up LP. The median number of days between the last rapamycin dose and follow-up lumbar puncture were 6 (sd = 5.12). The median duration between the baseline assessment of MoCA and first dose of rapamycin was 102 days. The follow-up cognitive assessment was performed within 28 days after the last dose of the study drug (median = 5 days).

The median duration between the baseline assessment of cognitive outcomes other than MoCA and first dose of rapamycin was 7.6 (range = 0.7 - 19.4) months. The follow-up cognitive assessment was performed within 28 days after the last dose of the study drug (median = 5 days, range = 0 - 19). The reason for the long duration between baseline cognitive assessment and study start was that all cognitive scores (except MoCA) were taken from the clinical work up of participants.

#### Supplementary Information 2

Cerebellar segmentation was performed using CerebNet [1], followed by morphological erosion (one voxel) of the cerebellar gray matter. Both vermis and cerebral cortex were dilated by four voxels. Overlapping parts between the dilated regions and the eroded cerebellum were removed to avoid PET signal spilling in from those regions. The result was a conservatively delineated gray matter cerebellum mask.

#### sFigure 1.

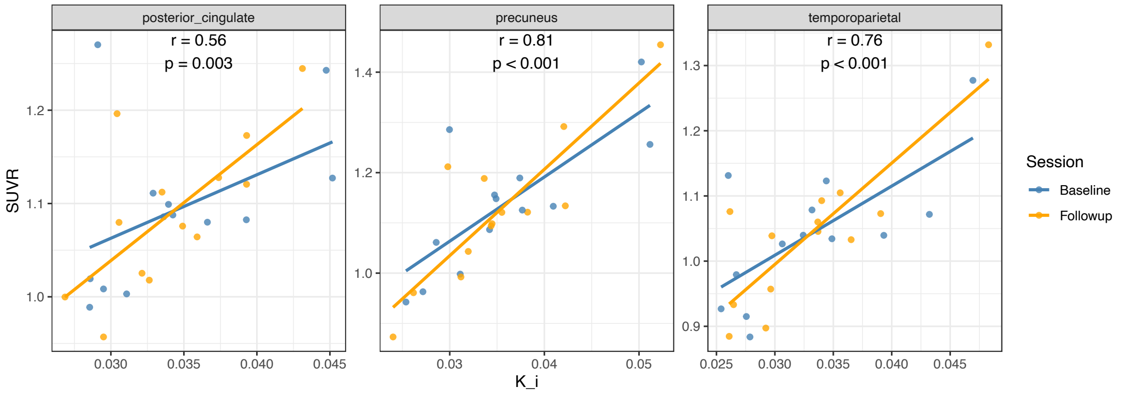


*sFigure 1. Correlations between [18]FDG $K_i$ and SUVR.*

#### sTable 1. Change in [18F]FDG SUVR

| Region | Primary Exploratory | Baseline Mean±SD | Followup Mean±SD | Change Mean±SD | Mean Change 95% CI | Percent Change Mean ± SD | t | p | Cohen's d |
|---|---|---|---|---|---|---|---|---|---|
| precuneus | Primary | 1.14±0.13 | 1.12±0.15 | -0.014±0.034 | [-0.035 0.007] | -1.34±3.01 | -1.45 | 0.17 | -0.40 |
| PCC | Primary | 1.09±0.08 | 1.07±0.08 | -0.010±0.030 | [-0.028 0.008] | -0.95±2.63 | -1.26 | 0.232 | -0.35 |
| temporoparietal | Primary | 1.04±0.10 | 1.04±0.11 | 0.000±0.031 | [-0.019 0.019] | -0.03±2.86 | 0.01 | 0.99 | 0.00 |
| total GM | Exploratory | 1.08±0.09 | 1.09±0.08 | 0.005±0.030 | [-0.013 0.023] | 0.55±2.63 | 0.66 | 0.52 | 0.18 |
| hippocampus | Exploratory | 0.73±0.06 | 0.74±0.05 | 0.015±0.028 | [-0.002 0.032] | 2.29±3.84 | 1.97 | 0.07 | 0.54 |
| amygdala | Exploratory | 0.70±0.04 | 0.72±0.03 | 0.016±0.031 | [-0.003 0.035] | 2.42±4.46 | 1.84 | 0.09 | 0.51 |
| PHCG | Exploratory | 0.88±0.06 | 0.89±0.06 | 0.012±0.028 | [-0.005 0.029] | 1.49±3.13 | 1.59 | 0.14 | 0.44 |
| insula | Exploratory | 0.92±0.05 | 0.93±0.05 | 0.018±0.025 | [0.003 0.034] | 2.07±2.80 | 2.58 | 0.02* | 0.72 |
| occipital | Exploratory | 1.12±0.11 | 1.14±0.11 | 0.014±0.050 | [-0.016 0.045] | 1.37±4.42 | 1.03 | 0.32 | 0.29 |
| frontal | Exploratory | 1.19±0.10 | 1.19±0.09 | 0.002±0.037 | [-0.020 0.025] | 0.28±3.00 | 0.23 | 0.82 | 0.06 |
| ACC | Exploratory | 0.96±0.07 | 0.98±0.06 | 0.021±0.024 | [0.007 0.036] | 2.31±2.50 | 3.20 | 0.008* | 0.88 |
| thalamus | Exploratory | 1.02±0.07 | 1.02±0.06 | 0.002±0.031 | [-0.017 0.021] | 0.28±3.01 | 0.22 | 0.83 | 0.06 |
| putamen | Exploratory | 1.24±0.09 | 1.27±0.08 | 0.030±0.032 | [0.011 0.050] | 2.52±2.78 | 3.36 | 0.006* | 0.93 |
| caudate | Exploratory | 1.08±0.11 | 1.10±0.11 | 0.019±0.046 | [-0.008 0.047] | 1.86±4.22 | 1.53 | 0.15 | 0.42 |

GM = greymatter; PHCG = parahippocampal gyrus; PCC = posterior cingulate cortex ; ACC = anterior cingulate cortex

**sTable 2. Change in [18F]FDG Ki**

| Region | Primary Exploratory | Baseline Mean±SD | Followup Mean±SD | Change Mean±SD | Mean Change 95% CI | Percent Change Mean ± SD | t | p | Cohen's d |
|---|---|---|---|---|---|---|---|---|---|
| precuneus | Primary | 0.036±0.008 | 0.035±0.007 | -6e-04±0.0038 | [-0.0029 0.0017] | -1.00±9.67 | -0.55 | 0.59 | -0.15 |
| PCC | Primary | 0.034±0.006 | 0.034±0.005 | 0.0006±0.004 | [-0.0029 0.0018] | -0.69±10.40 | -0.51 | 0.622 | -0.14 |
| temporoparietal | Primary | 0.033±0.007 | 0.033±0.006 | 0.0000±0.0031 | [-0.0019 0.0019] | 0.57±8.59 | -0.04 | 0.97 | -0.01 |
| total GM | Exploratory | 0.034±0.006 | 0.035±0.005 | 3e-04±0.0031 | [-0.0016 0.0022] | 1.56±8.66 | 0.37 | 0.72 | 0.10 |
| hippocampus | Exploratory | 0.022±0.004 | 0.023±0.004 | 8e-04±0.0024 | [-7e-04 0.0023] | 4.25±10.46 | 1.21 | 0.25 | 0.34 |
| amygdala | Exploratory | 0.021±0.003 | 0.021±0.003 | -2e-04±0.0017 | [-0.0012 8e-04] | -0.56±7.84 | -0.40 | 0.70 | -0.11 |
| PHCG | Exploratory | 0.027±0.004 | 0.027±0.004 | 2e-04±0.0021 | [-0.0011 0.0015] | 1.05±7.14 | 0.32 | 0.75 | 0.09 |
| insula | Exploratory | 0.029±0.004 | 0.029±0.003 | 7e-04±0.0023 | [-7e-04 0.0022] | 3.02±8.09 | 1.16 | 0.27 | 0.32 |
| occipital | Exploratory | 0.034±0.006 | 0.035±0.006 | 9e-04±0.0034 | [-0.0012 0.0029] | 3.35±9.58 | 0.93 | 0.37 | 0.26 |
| frontal | Exploratory | 0.038±0.006 | 0.039±0.005 | 4e-04±0.0037 | [-0.0019 0.0026] | 1.59±9.47 | 0.36 | 0.72 | 0.10 |
| ACC | Exploratory | 0.030±0.004 | 0.031±0.004 | 0.0011±0.0031 | [-0.0008 0.0029] | 3.94±9.60 | 1.28 | 0.23 | 0.35 |
| thalamus | Exploratory | 0.031±0.006 | 0.031±0.005 | 4e-04±0.0046 | [-0.0024 0.0032] | 2.50±14.93 | 0.29 | 0.78 | 0.08 |
| putamen | Exploratory | 0.040±0.007 | 0.042±0.005 | 0.0012±0.0040 | [-0.0012 0.0037] | 4.01±9.92 | 1.10 | 0.29 | 0.31 |
| caudate | Exploratory | 0.036±0.007 | 0.036±0.007 | 8e-04±0.0028 | [-9e-04 0.0024] | 2.51±7.33 | 1.02 | 0.33 | 0.28 |

GM = greymatter; PHCG = parahippocampal gyrus; PCC = posterior cingulate cortex ; ACC = anterior cingulate cortex

**sTable 3. Change in ASL MRI PLD 1.25s**

| Region | Primary Exploratory | Baseline Mean±SD | Followup Mean±SD | Change Mean±SD | Mean Change 95% CI | Percent Change Mean ± SD | t | p | Cohen's d |
|---|---|---|---|---|---|---|---|---|---|
| precuneus | Primary | 37.22±14.74 | 37.67±15.96 | 0.46±8.27 | [-4.54 5.46] | 2.27±28.92 | 0.20 | 0.85 | 0.06 |
| PCC | Primary | 47.50±11.50 | 46.80±14.90 | -0.73±8.33 | [-5.76 4.30] | -2.03±19.80 | -0.32 | 0.756 | -0.09 |
| temporoparietal | Primary | 41.47±12.63 | 41.66±13.39 | 0.19±6.60 | [-3.80 4.18] | 1.06±18.42 | 0.11 | 0.92 | 0.03 |
| total GM | Exploratory | 39.73±10.77 | 40.63±12.33 | 0.90±6.57 | [-3.07 4.87] | 2.49±18.05 | 0.49 | 0.63 | 0.14 |
| hippocampus | Exploratory | 47.58±11.99 | 48.61±12.63 | 1.04±6.41 | [-2.84 4.91] | 2.70±15.16 | 0.58 | 0.57 | 0.16 |
| amygdala | Exploratory | 42.53±8.57 | 44.79±10.57 | 2.26±6.81 | [-1.85 6.38] | 5.89±16.84 | 1.20 | 0.25 | 0.33 |
| PHCG | Exploratory | 36.73±12.22 | 39.05±13.36 | 2.33±6.85 | [-1.81 6.47] | 7.35±20.08 | 1.23 | 0.24 | 0.34 |
| insula | Exploratory | 50.52±7.09 | 49.81±9.06 | -0.71±4.45 | [-3.40 1.98] | -1.60±9.19 | -0.58 | 0.58 | -0.16 |
| occipital | Exploratory | 33.73±16.48 | 36.80±18.12 | 3.07±7.33 | [-1.35 7.50] | 10.45±22.15 | 1.51 | 0.16 | 0.42 |
| frontal | Exploratory | 41.82±9.55 | 42.20±11.50 | 0.38±6.79 | [-3.72 4.48] | 1.14±17.42 | 0.20 | 0.84 | 0.06 |
| ACC | Exploratory | 54.50±6.80 | 53.00±9.12 | -1.53±7.52 | [-6.07 3.02] | -2.43±14.6 | -0.73 | 0.48 | -0.20 |
| thalamus | Exploratory | 45.82±8.33 | 46.86±13.94 | 1.04±10.27 | [-5.16 7.25] | 2.09±24.45 | 0.37 | 0.72 | 0.10 |
| putamen | Exploratory | 47.63±5.01 | 48.36±6.99 | 0.73±6.39 | [-3.13 4.59] | 1.89±12.39 | 0.41 | 0.69 | 0.12 |
| caudate | Exploratory | 37.55±5.32 | 37.31±8.17 | -0.23±6.89 | [-4.40 3.93] | -0.16±17.95 | -0.12 | 0.91 | -0.03 |

GM = greymatter; PHCG = parahippocampal gyrus; PCC = posterior cingulate cortex ; ACC = anterior cingulate cortex

**sTable 4. Change in ASL MRI PLD 2.25s**

| Region | Primary Exploratory | Baseline Mean±SD | Followup Mean±SD | Change Mean±SD | Mean Change 95% CI | Percent Change Mean ± SD | t | p | Cohen's d |
|---|---|---|---|---|---|---|---|---|---|
| precuneus | Primary | 52.48±11.51 | 54.81±12.39 | 2.33±6.00 | [-1.30 5.96] | 4.79±11.96 | 1.40 | 0.19 | 0.39 |
| PCC | Primary | 54.70±9.69 | 56.5±10.1 | 1.80±5.02 | [-1.23 4.83] | 3.73±9.37 | 1.29 | 0.22 | 0.36 |
| temporoparietal | Primary | 52.00±11.12 | 52.39±10.89 | 0.39±6.17 | [-3.34 4.12] | 1.63±12.73 | 0.23 | 0.82 | 0.06 |
| total GM | Exploratory | 50.50±9.42 | 51.84±8.92 | 1.34±5.89 | [-2.22 4.90] | 3.55±12.54 | 0.82 | 0.43 | 0.23 |
| hippocampus | Exploratory | 47.98±12.82 | 50.28±9.73 | 2.30±6.71 | [-1.76 6.36] | 7.06±15.11 | 1.23 | 0.24 | 0.34 |
| amygdala | Exploratory | 41.79±10.76 | 44.46±10.20 | 2.66±9.75 | [-3.23 8.55] | 9.45±25.36 | 0.98 | 0.34 | 0.27 |
| PHCG | Exploratory | 45.70±11.14 | 49.05±9.65 | 3.35±7.24 | [-1.03 7.72] | 9.49±17.56 | 1.67 | 0.12 | 0.46 |
| insula | Exploratory | 50.21±8.90 | 51.19±9.16 | 0.99±5.32 | [-2.23 4.20] | 2.56±11.63 | 0.67 | 0.52 | 0.19 |
| occipital | Exploratory | 54.20±13.90 | 58.82±11.74 | 4.62±8.98 | [-0.81 10.04] | 11.12±20.54 | 1.85 | 0.09 | 0.51 |
| frontal | Exploratory | 51.62±7.83 | 52.45±7.84 | 0.83±5.37 | [-2.42 4.08] | 2.23±11.39 | 0.56 | 0.59 | 0.15 |
| ACC | Exploratory | 54.1±8.62 | 55.3±7.84 | 1.22±6.65 | [-2.80 5.24] | 3.29±13.6 | 0.66 | 0.52 | 0.18 |
| thalamus | Exploratory | 48.05±7.42 | 51.26±6.47 | 3.22±4.40 | [0.56 5.87] | 7.49±9.71 | 2.64 | 0.02* | 0.73 |
| putamen | Exploratory | 39.91±5.98 | 39.91±5.57 | 0.00±2.67 | [-1.61 1.62] | 0.40±7.37 | 0.01 | 1.00 | 0.00 |
| caudate | Exploratory | 34.88±7.14 | 35.23±5.74 | 0.35±4.03 | [-2.09 2.78] | 2.30±12.88 | 0.31 | 0.76 | 0.09 |

GM = greymatter; PHCG = parahippocampal gyrus; PCC = posterior cingulate cortex ; ACC = anterior cingulate cortex

#### sTable 5. Volumetric change in all regions

| Region | Primary Exploratory | Baseline Mean±SD | Followup Mean±SD | Change Mean±SD | Mean Change 95% CI | Percent Change Mean ± SD | t | p | Cohen's d |
|---|---|---|---|---|---|---|---|---|---|
| precuneus | Primary | 17763±3023 | 17458±2948 | -305±486 | [-599 -12] | -1.65±2.87 | -2.26 | 0.04* | -0.63 |
| PCC | Primary | 5370±788 | 5292±734 | -78±119 | [ -151 -6 ] | -1.32±2.35 | -2.37 | 0.04* | -0.66 |
| temporoparietal | Primary | 8534±11815 | 83832±12226 | -1502±1417 | [-2358 -646] | -1.84±1.93 | -3.82 | 0.002* | -1.06 |
| total GM | Exploratory | 487059±54274 | 481309±53753 | -5750±7086 | [-10032 -1468] | -1.17±1.52 | -2.93 | 0.01* | -0.81 |
| hippocampus | Exploratory | 7489±1120 | 7404±1140 | -85±136 | [-167 -2] | -1.17±1.96 | -2.24 | 0.04* | -0.62 |
| amygdala | Exploratory | 2987±547 | 2964±533 | -22±76 | [-68 23] | -0.70±2.39 | -1.07 | 0.31 | -0.30 |
| PHCG | Exploratory | 32977±5202 | 32540±4999 | -437±951 | [-1011 137] | -1.19±3.02 | -1.66 | 0.12 | -0.46 |
| insula | Exploratory | 13604±1139 | 13367±1357 | -238±586 | [-592 116] | -1.81±4.05 | -1.46 | 0.17 | -0.41 |
| occipital | Exploratory | 48774±8017 | 48120±7692 | -653±1068 | [-1299 -8] | -1.26±1.89 | -2.21 | 0.05* | -0.61 |
| frontal | Exploratory | 156206±15306 | 154843±15040 | -1363±3303 | [-3359 633] | -0.84±2.23 | -1.49 | 0.16 | -0.41 |
| ACC | Exploratory | 3895±555 | 3892±536 | -3±94 | [-59 53] | 0.04±2.68 | -0.11 | 0.91 | -0.03 |
| thalamus | Exploratory | 13374±1311 | 13314±1381 | -61±319 | [-253 132] | -0.48±2.23 | -0.69 | 0.51 | -0.19 |
| putamen | Exploratory | 9283±916 | 9193±872 | -91±121 | [-164 -17] | -0.94±1.23 | -2.70 | 0.02* | -0.75 |
| caudate | Exploratory | 6772±549 | 6755±596 | -17±122 | [-90 57] | -0.29±1.77 | -0.49 | 0.63 | -0.14 |

GM = greymatter; PHCG = parahippocampal gyrus; PCC = posterior cingulate cortex ; ACC = anterior cingulate cortex

#### sTable 6. CSF markers change

| Marker | N | Baseline Mean±SD | Followup Mean±SD | Change Mean±SD | Mean Change 95% CI | Percent Change Mean ± SD | t | p | Cohen's d |
|---|---|---|---|---|---|---|---|---|---|
| Aβ42 | 10 | 614±191 | 654±135 | 40±86 | [-21.76 101.56] | 9.91±15.18 | 1.46 | 0.177 | 0.46 |
| Aβ40 | 10 | 13621±3976 | 15137±4386 | 1516±1354 | [547.41 2484.66] | 11.96±11.87 | 3.54 | 0.006* | 1.12 |
| Aβ42/40 | 10 | 0.05±0.01 | 0.04±0.01 | 0.00±0.01 | [-0.01 0.001] | -1.49±12.26 | -0.59 | 0.571 | -0.19 |
| p181_tau | 10 | 107.1±34.4 | 110.1±33.3 | 3.0±17.0 | [-9.13 15.13] | 4.47±16.91 | 0.56 | 0.589 | 0.18 |
| Total tau | 10 | 616±150 | 751±206 | 135±103 | [61.26 208.94] | 21.78±16.71 | 4.14 | 0.003* | 1.31 |
| p181 tau/Total tau | 10 | 0.17±0.02 | 0.15±0.02 | -0.03±0.02 | [-0.04 -0.01] | -14.07±7.96 | -4.91 | 0.001* | -1.55 |
| Neurofilament | 10 | 1054±254 | 1281±346 | 227±219 | [70.67 383.33] | 23.46±25.96 | 3.29 | 0.010* | 1.04 |
| Albumin CSF | 10 | 227.1±75.1 | 241.1±116.6 | 14.0±53.5 | [-24.27 52.27] | 3.56±15.22 | 0.83 | 0.429 | 0.26 |
| Albumin CSF/plasma | 10 | 5.44±1.87 | 5.87±2.90 | 0.43±1.36 | [-0.54 1.40] | 5.28±15.89 | 1 | 0.344 | 0.32 |

#### sFigure 2. CSF markers change

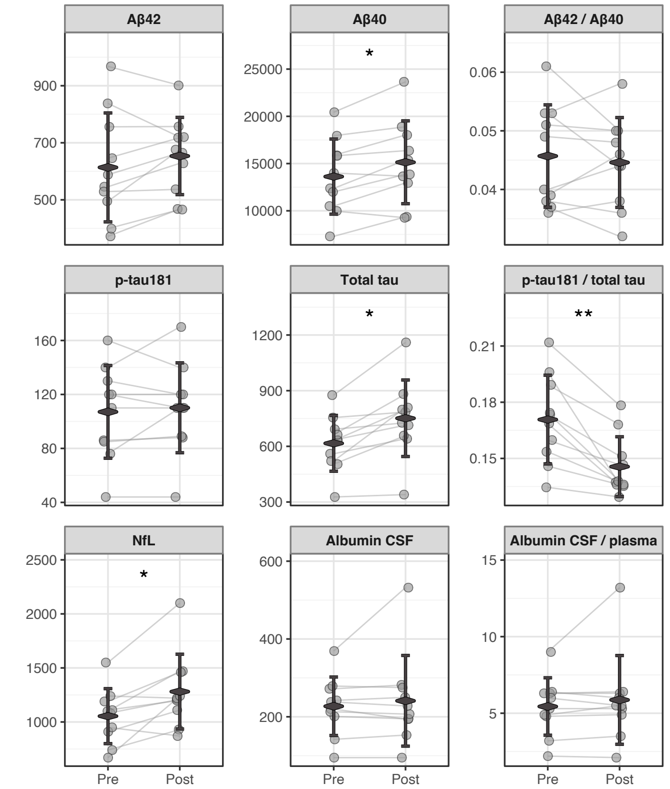


**p<0.01; **p<0.001*

**sTable 7. Change in CSF markers v.s. change in regional brain volume**

| CSF marker | Region | Pearson's r | p | df |
|---|---|---|---|---|
| AB40 | posterior cingulate | -0.34 | 0.34 | 8 |
| | precuneus | -0.19 | 0.60 | 8 |
| | temporoparietal | -0.11 | 0.76 | 8 |
| AB42 | posterior_cingulate | -0.14 | 0.69 | 8 |
| | precuneus | -0.24 | 0.50 | 8 |
| | temporoparietal | -0.54 | 0.11 | 8 |
| Neurofilament light | posterior_cingulate | 0.14 | 0.69 | 8 |
| | precuneus | 0.05 | 0.90 | 8 |
| | temporoparietal | -0.34 | 0.34 | 8 |
| total tau | posterior_cingulate | -0.04 | 0.92 | 8 |
| | precuneus | 0.25 | 0.48 | 8 |
| | temporoparietal | 0.56 | 0.09 | 8 |
| p-tau/total tau | posterior_cingulate | -0.42 | 0.23 | 8 |
| | precuneus | -0.17 | 0.65 | 8 |
| | temporoparietal | 0.25 | 0.48 | 8 |

**sTable 8. Change in cognitive score**

| Test | N | Baseline Mean±SD | Followup Mean±SD | Change Mean±SD | p |
|---|---|---|---|---|---|
| MoCA | 12 | 24.33 (2.93) | 24.75 (4.61) | 0.42 (2.31) | 0.55 |
| RCFT copy | 10 | 32.70 (2.36) | 29.20 (10.76) | -3.50 (10.37) | 0.31 |
| RCFT copy [s] | 8 | 249.88 (189.89) | 179.00 (71.55) | -70.88 (173.75) | 0.29 |
| RFCT immediate recall | 10 | 12.40 (6.98) | 11.20 (9.53) | -1.20 (7.42) | 0.62 |
| RAVLT learning | 10 | 35.50 (6.57) | 32.70 (9.19) | -2.80 (8.09) | 0.30 |
| RAVLT delay recall | 10 | 4.40 (3.66) | 4.70 (3.68) | 0.30 (3.20) | 0.77 |
| RAVLT recog true | 9 | 12.70 (2.06) | 13.10 (2.13) | 0.40 (2.37) | 0.61 |
| RAVLT recog false | 8 | 2.11 (2.26) | 3.56 (3.71) | 1.44 (3.61) | 0.26 |
| RAVLT recog corr | 8 | 10.33 (3.81) | 9.44 (5.15) | -0.89 (5.21) | 0.62 |
| Hagman test 1 | 10 | 13.80 (5.79) | 12.20 (6.14) | -1.60 (6.31) | 0.44 |
| Hagman test 2 | 10 | 17.30 (5.46) | 15.20 (6.65) | -2.10 (5.49) | 0.26 |

MoCA = Montreal Cognitive Assessment; RCFT = Rey-Osterrieth Complex Figure Test; RCFT copy = Rey-Osterrieth Complex Figure Test - Copy Trial; RCFT copy (s)= Rey-Osterrieth Complex Figure Test - Copy Time (seconds); RCFT immediate recall = Rey-Osterrieth Complex Figure Test - Immediate Recall; RAVLT = Rey Auditory Verbal Learning Test; RAVLT learning = Rey Auditory Verbal Learning Test - Learning Trials; RAVLT delay recall = Rey Auditory Verbal Learning Test - Delayed Recall; RAVLT recognition true = Rey Auditory Verbal Learning Test - Recognition Hits (True Positives); RAVLT recognition false = Rey Auditory Verbal Learning Test - Recognition False Positives; RAVLT recognition corr = Rey Auditory Verbal Learning Test - Corrected Recognition Score (Hits - False Positives).
